# Estimation of the test to test distribution as a proxy for generation interval distribution for the Omicron variant in England

**DOI:** 10.1101/2022.01.08.22268920

**Authors:** Sam Abbott, Katharine Sherratt, Moritz Gerstung, Sebastian Funk

## Abstract

**Background:** Early estimates from South Africa indicated that the Omicron COVID-19 variant may be both more transmissible and have greater immune escape than the previously dominant Delta variant. The rapid turnover of the latest epidemic wave in South Africa as well as initial evidence from contact tracing and household infection studies has prompted speculation that the generation time of the Omicron variant may be shorter in comparable settings than the generation time of the Delta variant.

**Methods:** We estimated daily growth rates for the Omicron and Delta variants in each UKHSA region from the 23rd of November to the 23rd of December 2021 using surveillance case counts by date of specimen and S-gene target failure status with an autoregressive model that allowed for time-varying differences in the transmission advantage of the Delta variant where the evidence supported this. By assuming a gamma distributed generation distribution we then estimated the generation time distribution and transmission advantage of the Omicron variant that would be required to explain this time varying advantage. We repeated this estimation process using two different prior estimates for the generation time of the Delta variant first based on household transmission and then based on its intrinsic generation time.

**Results:** Visualising our growth rate estimates provided initial evidence for a difference in generation time distributions. Assuming a generation time distribution for Delta with a mean of 2.5-4 days (90% credible interval) and a standard deviation of 1.9-3 days we estimated a shorter generation time distribution for Omicron with a mean of 1.5-3.2 days and a standard deviation of 1.3-4.6 days. This implied a transmission advantage for Omicron in this setting of 160%-210% compared to Delta. We found similar relative results using an estimate of the intrinsic generation time for Delta though all estimates increased in magnitude due to the longer assumed generation time.

**Conclusions:** We found that a reduction in the generation time of Omicron compared to Delta was able to explain the observed variation over time in the transmission advantage of the Omicron variant. However, this analysis cannot rule out the role of other factors such as differences in the populations the variants were mixing in, differences in immune escape between variants or bias due to using the test to test distribution as a proxy for the generation time distribution.

## Introduction

Early estimates from South Africa indicated that the Omicron COVID-19 variant may be both more transmissible and have greater immune escape than the previously dominant Delta variant^[2]^. The rapid turnover of the latest epidemic wave in South Africa has prompted speculation that the generation time of the Omicron variant may be shorter in comparable settings than the generation time of the Delta variant. Recent observations of serial intervals in South Korea^[3]^ supports this hypothesis though with a large degree of uncertainty and a small sample size. Other recent studies have indicated that Omicron may have a shorter incubation period than Delta, adding some potential support for this theory^[4]^. However, the relationship between symptom presentation, immunity (and potential earlier developoment of symptoms in the presence thereof), contact behaviour (potentially modulated by self-isolation following symptoms) and generation times is far from trivial.

Previously^[5]^, we estimated growth rates for the Omicron and Delta variants jointly in England by UKHSA region. We used S-gene target failure (SGTF) as a proxy of variant status (with SGTF highly correlated with Omicron) combined with reported case counts. A key finding from this report was that the growth advantage of Omicron variant versus the Delta variant appeared to vary over time. There are a range of potential explanations for this finding, such as: the impact of differential immune escape, sampling bias in the SGTF data, differences in the populations in which the variants are circulating (for example differences in age group mixtures), the impact of behavioural or policy change, or differing generation times between variants in comparable settings.

In this study, we focus on estimating the generation time distribution that would be required to explain the observed variation in growth advantage. We do so by exploiting the approximate relationship between the daily growth rate, gamma distributed generation times, and the effective reproduction number^[6,7]^. This relationship means that estimates of the growth rate for the Delta variant can be combined with prior estimates for the generation time of the Delta variant in a model in order to estimate the generation time of Omicron based on the observed variation in transmission advantage between the Omicron and Delta variant. We use growth rate estimates by UKHSA region derived using surveillance case counts and SGTF data along with prior estimates for the generation time from Hart et al^[8]^.

## Methods

### Data

We use test-positive cases’ reported S-gene status as a direct proxy for the Omicron variant, with a target failure indicating a case has the Omicron variant. We augment this data with total reported cases counts. Both sets of data were sourced from UKHSA, available daily by specimen date for England and by region. We use data from the 23rd of November 2021 up to the 23rd of December 2021. This covers the main transition period from the Delta variant being dominant to the Omicron variant being dominant and exludes the majority of the Christmas reporting period in which test seeking behaviour became atypical.

### Growth rate estimation

We use the same methodology to estimate variant specific growth rates as in our earlier study estimating the time-varying transmission advantage of the Omicron variant (archived here)^[5]^. Briefly, we use an autoregressive (AR) approach where expected cases for each variant are modelled as a combination of expected cases from the previous day and the exponential of the log growth rate for that variant and time point. The variant specific growth rate is then itself modelled as a differenced AR(1) process using a vector autoregression structure, which assumes that the first differences of the variant growth rates are drawn from a multivariate normal distribution. This model formulation can account for variant differences other than a transmission advantage in a principled manner. Crucially, in the absence of evidence that variants do not co-vary, it reduces to a model with a fixed transmission advantage.

We fit jointly to reported cases and SGTF data for each timeseries independently assuming a negative binomial and beta-binomial observation model respectively. Day of the week reporting periodicity for case counts is captured using a random effect for the day of the week. We initialise the model by fitting to a week of case-only data where the Omicron variant is assumed to not be present (from the 17th of November to the 22nd).

A full description of this model can be found in the documentation for the `forecast.vocs R` package^[9]^.

### Generation time and transmission advantage estimation

We model the relationship between estimated variant growth rates, their generation times, and the between variant transmission advantage using the gamma approximation framework developed by Park et al.^[6,7]^ to estimate the effective reproduction number for each variant assuming that their generation time is gamma distributed. This approximation is as follows and holds when growth is approximately exponential,

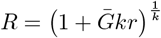

Where *R* is the effective reproduction number, 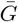 is the mean of the gamma distributed generation time, *r* is the daily growth rate, and *k* is the squared coefficient of variation which can be reparametrised as 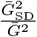 where 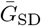 is the standard deviation of the gamma distribution.

Using this approximation we first estimate the effective reproduction number (*R*^D^) of the Delta variant across locations (*l*) and times (*t*, in days) using estimates of the daily growth rates (*r*^D^), and a gamma distributed generation time. Under an assumption of homogeneous mixing we can then relate the effective reproduction number of Delta to the effective reproduction number of Omicron (*R*^O^) by assuming a transmission advantage (*α*) which corresponds to a combination of any intrinsic transmission advantage as well as population based advantages (such as advantage gained from immune escape). Finally, we can then reverse the gamma approximation framework to estimate the daily growth rates for Omicron (*r*^O^) and compare these to those estimated directly in the previous modelling step.

Our model is described by the following set of equations:

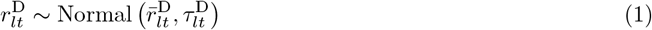

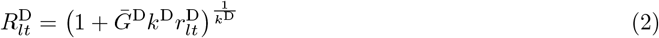

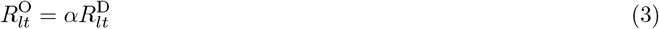

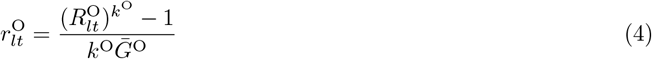

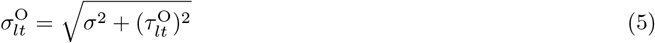

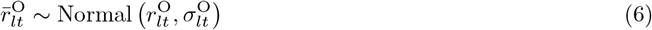

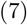

Here, superscripts D and O denote Delta and Omicron, respectively, *r* is the growth rate, 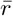 is the mean estimated growth rate from the previous modelling step, *τ* its standard deviation and *R* the effective reproduction number. We assume that generation time posteriors from the previous step are distributed according to independent normal distribtions and include an additional observation error (*σ*) to account for the approximate nature of the relationship between variant growth rates. The relationship between generation time estimates for Omicron and Delta is then modelled as follows,

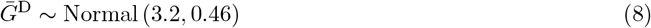

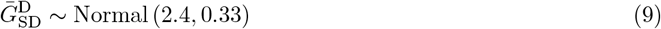

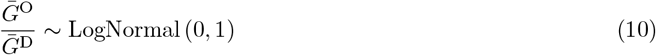

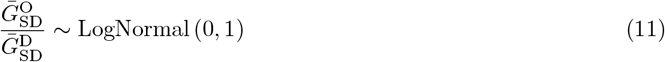

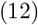

The generation priors used here are motivated by the household estimates derived in Hart et al^[8]^ for the UK using a normal approximation. As a sensitivity analysis, we also explore their alternative intrinsic estimates which had a mean of 4.6 days (standard deviation (SD): 0.36 days) and a standard deviation of 3.1 days (SD: 0.18) days, again using a normal approximation. Finally we assume that the standard deviation of the reporting delay has a normal prior truncated at zero with a mean of zero and a standard deviation of 0.1.

### Statistical inference

We first visualised our combined data sources (cases by specimen date and SGTF status by specimen date). We then fit the growth rate estimation model separately to data for each UKHSA region and extracted posterior estimates of the growth rate for both Omicron and Delta, and overall. To motivate subsequent modelling we then visualised these estimates over time and in comparison to each other. We added linear fits to the growth rate comparison plot for each UKHSA region as well as overall in order to visualise the relationship between variants compared to the idealised relationship in which they shared a generation time estimate. This was motivated by the simple effective reproduction number approximation 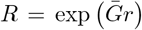. This means that the gradient of the relationship between variant growth rates can be considered as the approximate ratio between variant generation time means (and so should be 1 when they share the same generation time).

We then used the mean and standard deviation of the growth rate estimates from the 1st to the 23rd December 2021, excluding early growth estimates due to the potential for bias from imported cases. These were generated by the first modelling step to jointly estimate the generation time distribution for Omicron as well as its overall transmission advantage compared to Delta. This assumes that the generation time distribution for both variants as well as the transmission advantage is the same over all UKHSA regions and across time. We used both the household and intrinsic Delta generation time estimates from Hart et al.^[8]^ and presented the resulting posterior estimates from both. To check the performance of our modelling approach, we plot the posterior predictions for Omicron daily growth rates by UKHSA region over time and compared them to the estimates from the growth rate estimation step.

### Implementation

The growth rate estimation model was implemented using the `forecast.vocs R` package^[9,10]^ and fit using `stan`^[11]^ and `cmdstanr`^[12]^. The model was fit using 2 chains with each chain having 1000 warmup steps and 2000 sampling steps. The generation time estimation model was also implemented using `stan`^[11]^ and `cmdstanr`^[12]^ with the default of 4 chains each with 1000 warmup and 1000 sampling steps. For all models convergence was assessed using the Rhat diagnostic^[11]^ and fitting was initialised using samples drawn from near the mean of each prior distributions.

## Results

### Data description

**Growth rate of reported cases overall, with the Omicron variant, and not with the Omicron variant**

**Over time**

**Omicron versus Delta**

**Generation time and transmission advantage estimates**

**Table 1:**
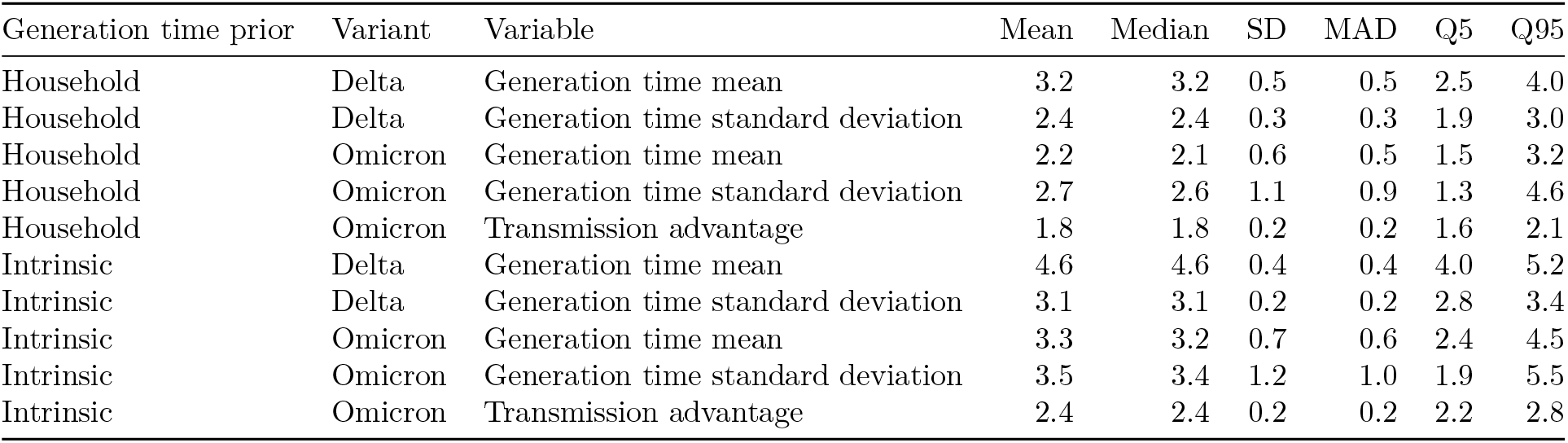
Posterior estimates for generation time and transmission advantage by choice of generation time prior

**Posterior predictions**

## Discussion

We found that a shorter generation time was consistent with the time-varying transmission advantage observed for the Omicron variant in England from the beginning of December to the 23rd of December 2021.

**Figure 1:**
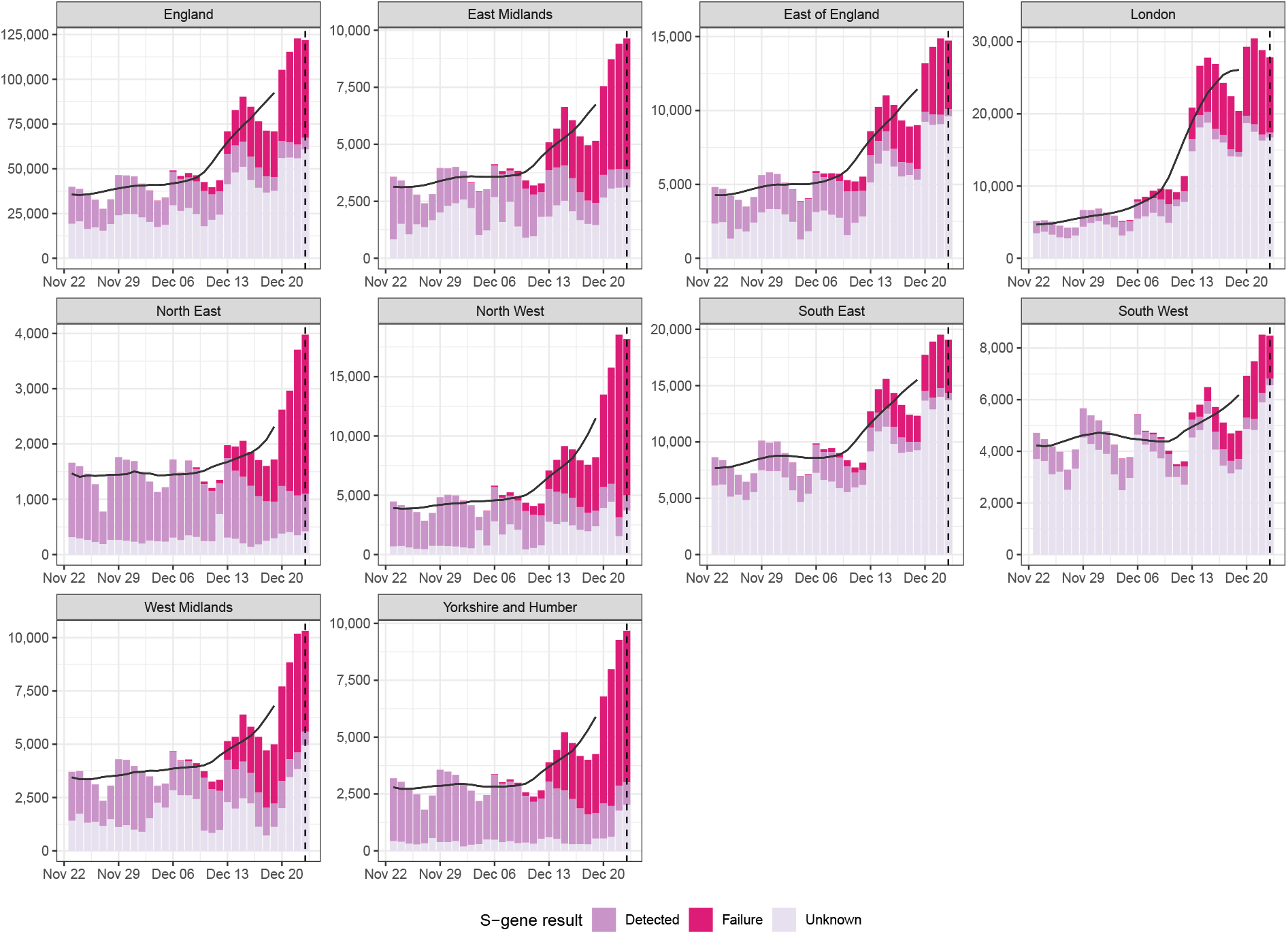
Daily cases in England and by UKHSA region, with S-gene target result (failed, confirmed detected, or unknown), and centred 7-day moving average starting from the 23rd of November 2021 and including data up to the 23rd of December 2021 (dotted line). Source: UKHSA and coronavirus.gov.uk; data by specimen date.

**Figure 2:**
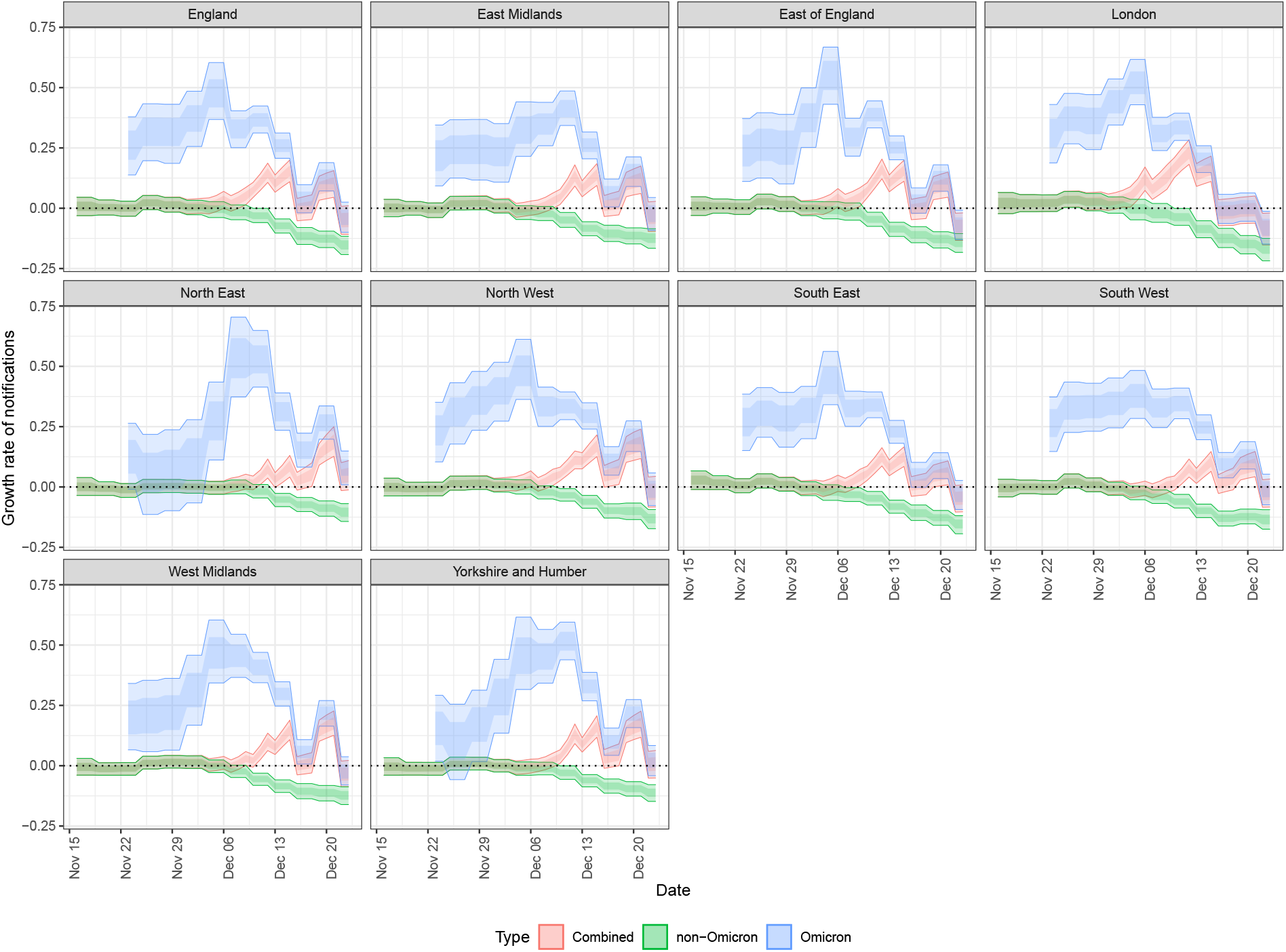
Estimates of the time-varying daily growth rate for England and by UKHSA region (using the log scale approximation) for reported cases overall, cases with the Omicron variant, and non-Omicron cases. Growth rates are assumed to be piecewise constant with potential changes every 3 days. Growth rates are assumed to be constant beyond the forecast horizon (vertical dashed line). 90% and 60% credible intervals are shown.

**Figure 3:**
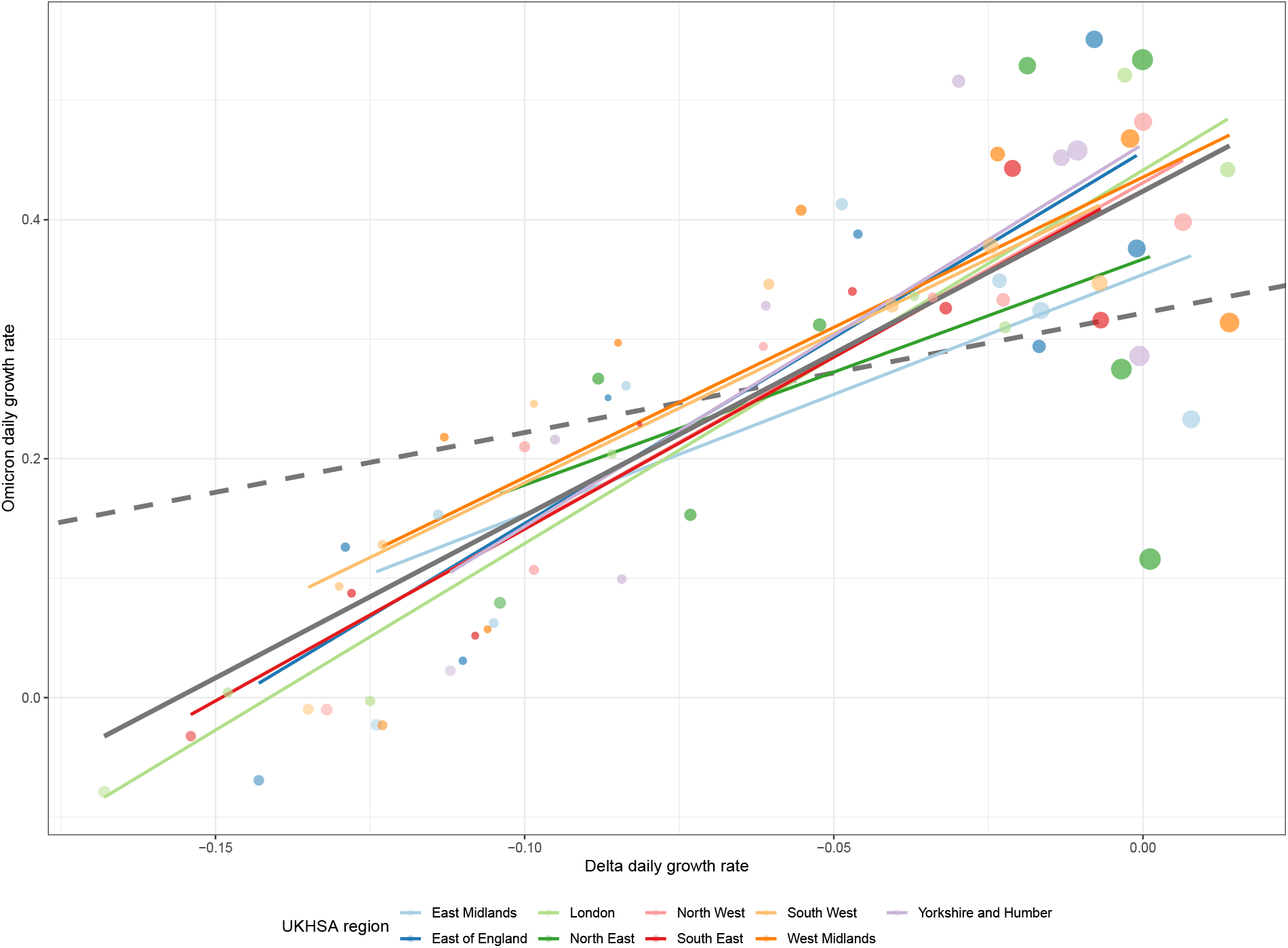
Estimates of the time-varying daily growth rate for each UKHSA region for cases with the Omicron variant, and non-Omicron cases. Point size has been used to represent the uncertainty of each estimate (though it does not correspond to a meaningful credible interval). The dashed grey line indicates the expected relationship between variants if they differ only by a fixed transmission advantage. The solid grey line indicates the observed overall linear relationship between variants, and the slope of this line approximates the ratio between generation time means. The region specific lines show how this relationship differs between regions.

**Figure 4:**
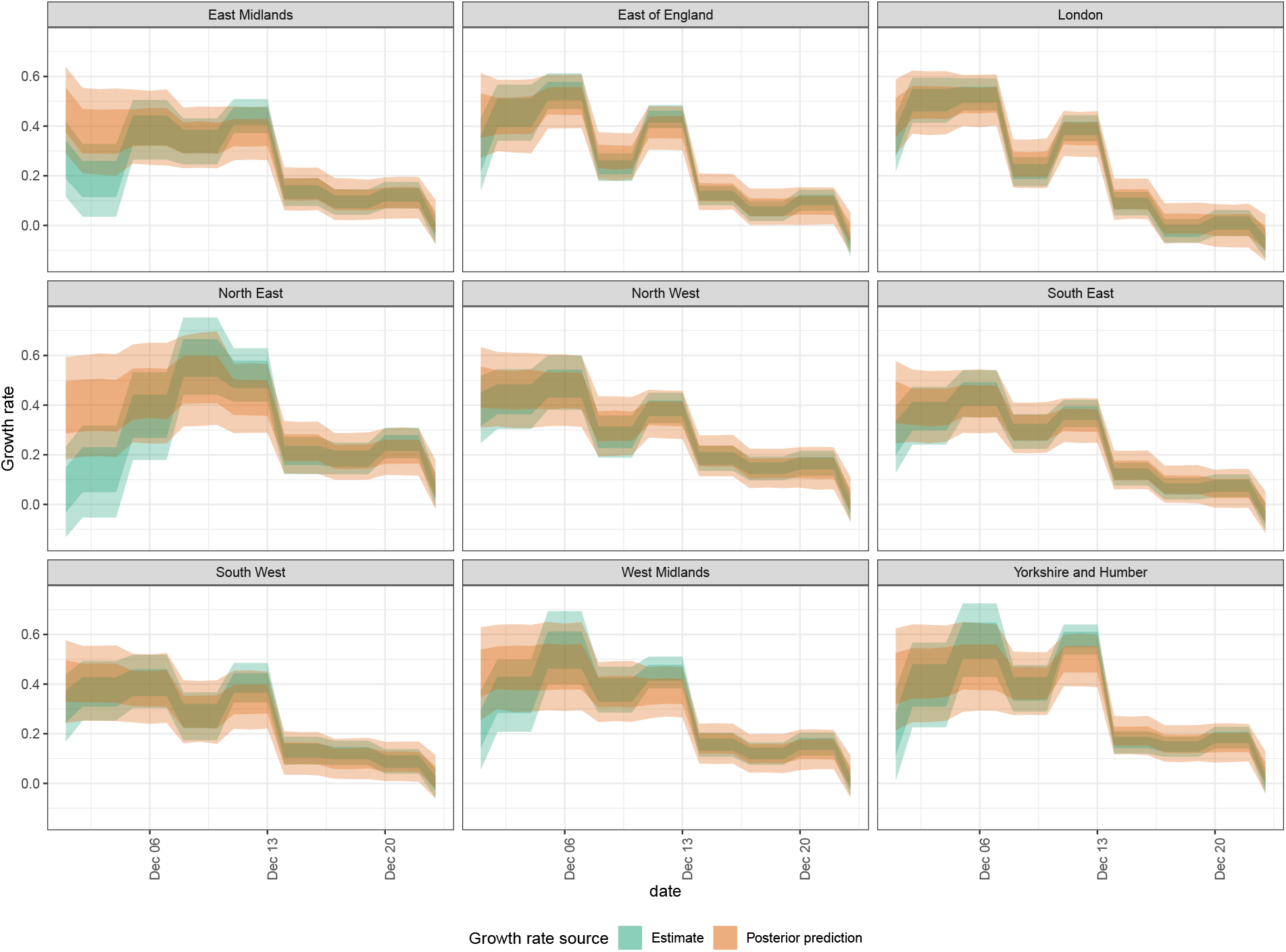
Estimates of the time-varying daily growth rate for each UKHSA region for cases with the Omicron variant compared to posterior predictions using Delta growth rates and differing generation times. Delta growth rates are assumed to be piecewise constant with potential changes every 3 days. 90% and 60% credible intervals are shown.

Assuming a generation time distribution for Delta with a mean of 2.5-4 days (90% credible interval) and a standard deviation of 1.9-3 days we estimated a shorter generation time distribution for Omicron, with a mean of 1.5-3.2 days and a standard deviation of 1.3-4.6 days. This implied a transmission advantage for Omicron in this setting of 160%-210% compared to Delta. We found similar relative results using an estimate of the intrinsic generation time for Delta, though all estimates increased in magnitude due to the longer assumed generation time. However, our analysis may be biased by a range of factors and so these findings should be considered in the light of other results.

Our analysis had several key limitations. Firstly, the surveillance data sources we used to estimate daily growth rates had the potential to bias our analysis both because SGTF may be an imperfect proxy for Omicron and because we used public case count data which do not include reinfections. We made no adjustment for the background rate of SGTF observed for Delta or other factors that might lead to SGTF, or a confirmed detected S-gene observed with Omicron. The exclusion of known reinfections could also lead to bias in our estimates if the proportion of infections that are reinfections varies over time and between variants. Changes in testing practice may also impact our results, especially if this occurs differentially in populations where different proportions of each variant are circulating. In our first study^[5]^, we explored the potential for sampling bias for cases with known S-gene status and found little evidence that this was present.

Our generation time and transmission advantage estimates for Omicron also have limitations. A major limitation is that our estimated generation time distributions are in fact the distributions from a positive test to resulting positive tests rather than the true generation time distribution which would be based on infection times. This means that our estimates may be biased if those secondary infections test positive prior to the original (primary) infected case, or may be biased by the epidemic phase^[13]^. In addition to this, our model only estimates an overall transmission advantage. We do not attempt to distinguish between “intrinsic” transmissibility and immune escape, a combination of which is likely to be underlying any sustained differences in growth rate. It is also possible that both the observed transmission advantage and generation times vary across UKHSA regions due to differences in population structure, prior immunity and other factors. However, our posterior predictive model checks imply a good match between observed and estimated growth rates for Omicron for the majority of data points indicating that this effect may be minimal. Finally, our estimates are at the national scale stratified by UKHSA region. There may be additional variation within this stratification that these estimates do not account for and this may bias our results.

There are several alternative explanations for our findings other than a change in the generation time distribution. One potential explanation is that that Omicron and Delta may have had differing age profiles in the time period studied, these age distributions may have changed over time, and age groups may have differing transmission potentials. This could potentially lead to the observed variation in transmission advantage between Omicron and Delta without requiring differing generation times. Similarly, if age groups change behaviour differently over time or respond differently to policy changes this could lead to similar changes in transmission advantage as observed.

Our results may also be the result of differential immune escape between variants. The reduction in the observed transmission advantage of Omicron over time could also be the result of Delta having greater immune escape from previous Omicron infection than Omicron. However, this seems unlikely given epidemiological evidence^[14]^ and the short time lines involved. As observed in Volz et al. social distancing typically reduces the number of people contacted per day, but increases the duration and frequency of household contacts^[13]^. This can result in the saturation of available contacts and hence reduce estimates of the transmission advantage for a variant. If social contacts changed markedly during our study period this could also be an explanation for the reduction over time in the observed transmission advantage of Omicron. Finally, the variants may have been circulating in populations that differed in more than just the distribution of ages. This would likely have a similar impact in terms of biasing our results.

Despite the range of potential sources of bias our results do correspond with observations of reduced serial intervals in South Korea^[3]^ and a potentially reduced incubation period^[4]^ although both these studies reported a large degree of uncertainty and the relationship between symptom presentation, immunity, contact behaviour and generation times is far from trivial.

Our results may also be the result of differential immune escape between variants. The reduction in the observed transmission advantage of Omicron over time could result from Delta having greater immune escape from previous Omicron infection than Omicron. However, this seems unlikely given epidemiological evidence^[14]^ and the short time lines involved. As observed in Volz et al. social distancing typically reduces the number of people contacted per day, but increases the duration and frequency of household contacts^[13]^. This can result in the saturation of available contacts and hence reduce estimates of the transmission advantage for a variant. If social contacts changed markedly during our study period this could be an explanation for the reduction over time in the observed transmission advantage of Omicron. Finally, the variants may have been circulating in populations that differed in more than just the distribution of ages. This would likely have a similar impact in terms of biasing our results. Despite the range of potential sources of bias our results do correspond with observations of reduced serial intervals in South Korea^[3]^ and a potentially reduced incubation period^[4]^, although both these studies reported a large degree of uncertainty and the relationship between symptom presentation, immunity, contact behaviour and generation times is far from trivial.

We found that a reduction in the generation time of Omicron compared to Delta was able to explain the observed variation over time in the transmission advantage of the Omicron variant. However, this analysis cannot rule out the role of other factors such as differences in the populations the variants were mixing in, differences in immune escape between variants, or a bias due to using the test-to-test distribution as a proxy for the generation time. More work is needed to explore these factors both using similar methods to those described in this study, potentially extending to a latent infection based model and/or stratifying by age, and also using alternative study designs such as household transmission studies and contact tracing studies.

## Data Availability

All data produced and code used are available online at https://github.com/epiforecasts/omicron-sgtf-forecast

